# Complete Loss of PAX4 causes Transient Neonatal Diabetes in Humans

**DOI:** 10.1101/2025.04.01.25324926

**Authors:** James Russ-Silsby, Yunkyeong Lee, Varsha Rajesh, Mahsa Amoli, Nasser Ali Mirhosseini, Tushar Godbole, Matthew B. Johnson, Dora E. Ibarra, Han Sun, Nicole A. J. Krentz, Matthew N. Wakeling, Sarah E. Flanagan, Andrew T. Hattersley, Anna L. Gloyn, Elisa De Franco

## Abstract

Gene discovery studies in individuals with diabetes diagnosed within 6 months of life (neonatal diabetes, NDM) can provide unique insights into the development and function of human pancreatic beta-cells. We describe the identification of homozygous *PAX4* loss-of-function variants in 2 unrelated individuals with NDM: a p.(Arg126*) stop-gain variant and a c.-352_104del deletion affecting the first 4 *PAX4* exons. We confirmed the p.(Arg126*) variant causes nonsense mediated decay in CRISPR-edited human induced pluripotent stem cell (iPSC)-derived pancreatic endoderm cells. Integrated analysis of CUT&RUN and RNA-sequencing in *PAX4*-depleted islet cell models identified genes directly regulated by PAX4 involved in both pancreatic islet development and glucose-stimulated insulin secretion. Both probands had transient NDM which remitted in early infancy but relapsed between the ages of 2 and 7 years, demonstrating that in contrast to mouse models, PAX4 is not essential for the development of human pancreatic beta-cells.

## 1. Introduction

The identification of genes in which bi-allelic loss-of-function (LoF) variants cause diabetes onset in the first 6 months of life (neonatal diabetes mellitus, NDM) can provide unique insights into the development and function of human pancreatic beta cells. Historically, animal gene knockout models, in particular mice, have been used to study pancreatic development. However, many differences exist between human and murine pancreatic development, including the transcription factors which control the process [1]. Biallelic LoF variants naturally mimic the functional consequences of gene knockouts examined in animal models. Thus, by studying individuals with recessive NDM, we can identify human-specific effects of losing genes involved in pancreatic development [2–4]. An example of this is the recent identification of biallelic LoF variants in *ZNF808* as a cause of NDM and pancreatic agenesis [5]. *ZNF808* is a primate-specific gene which is absent in all other mammals, including rodents. Through gene discovery in NDM, it has been demonstrated to have a critical role in pancreatic cell fate specification during human development.

In this study we use genome sequencing to identify homozygous *PAX4* LoF variants as a novel cause of NDM and functionally explore the human role of the gene using Cleavage Under Targets and Release Using Nuclease (CUT&RUN) [6] and RNA sequencing in human beta cell models.

## 2. Results and Discussion

### 2.1 Homozygous *PAX4* LoF variants are a novel cause of transient neonatal diabetes

Through genome sequencing, we identified homozygous LoF variants in the pancreatic homeobox transcription factor gene *PAX4* in two individuals. Both were diagnosed with transient neonatal diabetes (TNDM), a subtype of NDM that is characterised by a period of remission in infancy and childhood where no diabetes treatment is required (Figure 1). No other genes containing homozygous LoF variants were identified in more than one individual in the consanguineous NDM study cohort (N=43; 19 with TNDM), within whom all known genetic causes of NDM had previously been excluded through comprehensive genetic testing of the known NDM etiologies [7,8]. *PAX4* has long been considered a strong candidate gene for NDM due to its observed role in mammalian beta cell development [9–11] and its known association with type 2 diabetes [12]. The homozygous *PAX4* LoF variants identified were a p.(Arg126*) stop gain in individual 1 (I-1) and a 2.65kb deletion on chromosome 7 (c.-352_104del; GRCh37(chr7): g.127255495_127258142del) in individual 2 (I-2). The stop gain variant was heterozygous in 9 individuals in the gnomAD v4 database [13], which contains single variant and indel exonic data for 807,162 individuals, giving it an allele frequency of 5.56×10^-6^. The deletion was not present in the gnomAD v4 structural variant (SV) dataset that contains genome-wide structural variant data for 63,046 individuals. No high-confidence homozygous *PAX4* LoF variants were present in gnomAD v4, supporting biallelic loss-of-function variants being deleterious. Both variants were predicted to result in complete loss of *PAX4* mRNA: the stop gain variant p.(Arg126*) is located in the 6th of 12 exons of the Matched Annotation from NCBI and EMBL-EBI (MANE) select *PAX4* transcript (ENST00000639438) and is therefore predicted to result in mRNA degradation through nonsense-mediated decay (NMD), while the 2.65kb deletion (c.-352_104del) identified in I-2 removes the first 4 exons of the gene and its promoter.

**Figure 1:**
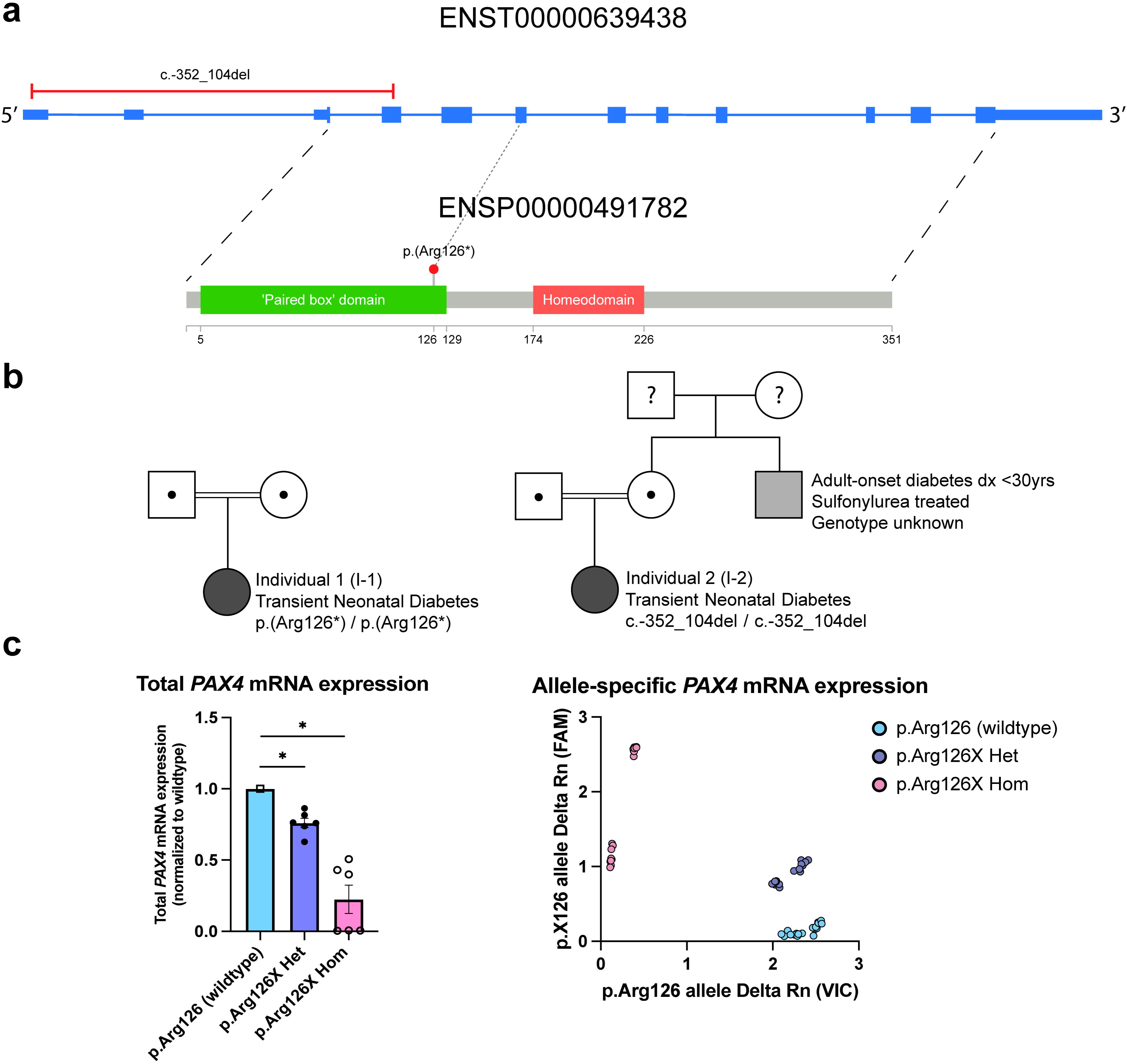
a) Two homozygous *PAX4* loss of function (LoF) variants were identified in two unrelated individuals. The variants are both predicted to cause complete ablation of PAX4 translation. b) The two individuals (filled black circle) had transient neonatal diabetes and were born to consanguineous parents. No additional individuals with TNDM were reported in either family. c) Reduction in *PAX4* transcript in iPSC-PE harboring p. (Arg126*) variant. A mean ∼24% reduction in total *PAX4* transcript expression was observed in the p.(Arg126*) heterozygous samples (two independent clones performed in triplicate) and a mean ∼78% reduction in the p.(Arg126*) homozygous samples (two independent clones performed in triplicate) when compared to the wildtype samples (two independent clones performed in triplicate). Allele-specific qPCR for the *PAX4* transcript corroborated the results.

We investigated the impact of the p.(Arg126*) variant on *PAX4* transcript levels by generating CRISPR-edited iPSCs harboring heterozygous and homozygous p.(Arg126)* variants. These iPSCs were differentiated into pancreatic endoderm cells, a stage where *PAX4* is abundantly expressed, following the protocol described by Rezania et al [14]. Total *PAX4* expression was then compared across isogenic controls (CRISPR-sham), heterozygous, and homozygous p.(Arg126*) cell lines. *PAX4* p.(Arg126*) heterozygous cells had a 24% reduction in transcript levels and homozygous cells had a 78% reduction in homozygous cells (Figure 1c). Allele-specific qPCR corroborated the abundance of *PAX4* transcript in relation to each genotype (Figure 1c).

We next investigated the presence of autosomal recessive *PAX4* variants in a replication cohort of 6,087 individuals with suspected monogenic diabetes using a combination of targeted next generation sequencing (tNGS) and genome sequencing. No additional homozygous or compound heterozygous *PAX4* LoF variants were identified. This replication cohort included 476 individuals with NDM diagnosed before 6 months and 5,611 individuals with clinically suspected monogenic diabetes (age at diagnosed range: 6 months to 60 years). 383 individuals were born to consanguineous parents. The absence of biallelic LoF variants in the replication cohort suggests that loss of *PAX4* is a rare cause of monogenic diabetes.

The two individuals with homozygous *PAX4* LoF variants had similar clinical features (Supplemental table 1). Both were diagnosed with diabetes between 1 and 5 months after birth. Diabetes remission occurred for both between 6 and 12 months of age and both relapsed between 2 and 7 years. Age at diabetes onset and remission are similar to those observed in individuals with TNDM caused by activating mutations in the K_ATP_ channel genes *ABCC8* and *KCNJ11*, which has a median age at onset of 4 weeks (range: 0-16 weeks) [15] and a median age upon entering remission of 8 months [15]. The ages at relapse are also consistent with the relapse range for K_ATP_-related TNDM, which typically occurs between 3-15 years (median: 4.7 years) [15]. Both children with *PAX4*-TNDM were treated with insulin at diagnosis and upon relapse (dose at last assessment for I-1 1U/Kg/day, dose not available for I-2). They also had reduced birth weights for their gestational age at -1.16 SD and -2.98 SD, consistent with reduced *in utero* insulin secretion during the third trimester, a critical period in foetal growth where insulin serves as the primary growth factor.[16] No extra pancreatic features were reported in either patient.

The parents of the two probands, who are heterozygous carriers of the respective variants, did not have diabetes at the time of recruitment. However, the parents of the proband with the p.(Arg126*) variants were both under the age of 35 (the age of the parents was not recorded for the second individual). This is younger than the age at diabetes onset for two of three individuals heterozygous for the previously reported p.(Tyr186*) *PAX4* LoF variant that was shown to be associated with increased risk of type 2 diabetes (T2D) in a single family [12]. The only reported family history of diabetes in either of our probands was in the maternal uncle of I-2, who was diagnosed with diabetes before the age of 30, is reported to be lean (BMI unknown), and is currently treated with a sulphonylurea. A DNA sample from this individual was not available for testing. The lack of a clear family history of diabetes in either family supports the finding from Laver *et al* [17] that *PAX4* heterozygous variants are unlikely to be a cause of maturity-onset diabetes of the young (MODY) but may confer an increased risk of type 2 diabetes, as was found by Lau *et al* [12].

### 2.2 PAX4 regulates pancreatic beta cell development and glucose sensitive insulin secretion

The identification of biallelic *PAX4* LoF variants as causing transient rather than permanent NDM is suggestive of a different role for PAX4 in human pancreatic development compared to that in the mouse. In mice, homozygous *Pax4* knockout (KO) results in a near complete lack of mature pancreatic beta and delta cells and a significantly elevated alpha cell count compared to wild type mice [9]. This in turn results in an inability of the murine pancreas to produce insulin or somatostatin, while glucagon production is significantly increased. *Pax4* KO mice are therefore born with diabetes and die shortly after birth. In contrast, the period of diabetes remission observed in our 2 patients with complete *PAX4* loss indicates that insulin-producing beta cells do still develop and are present in sufficient numbers for full glycaemic control during the remission period. Thus, our study provides another example of the differences in pancreatic development that exist between mice and humans [1]. Despite this, the observed transient remission suggests that PAX4 may contribute to beta cell proliferation, mirroring the ability of PAX4-expressing beta cells in mice to expand during pregnancy and periods of insulin resistance, and implying that certain aspects of PAX4-driven beta cell maintenance are conserved between species.[18]

To identify direct targets of PAX4 and explore its role in human beta cell development and function, we performed a CUT&RUN [6] assay to identify PAX4-bound regions in the EndoC-βH1 human pancreatic beta cell model.[19] Due to the unavailability of ChIP-grade PAX4 antibodies, we introduced FLAG-PAX4-V5-GFP via lentiviral transduction and sequenced 5 million reads from PAX4-V5 antibody-bound regions (Figure 2a). Analysis using SEACR [20] identified 1,673 binding peaks (Supplemental table 2). We then compared these peaks with chromatin accessibility data in adult donor beta cells from Chiou *et al* [21] and embryonic stem cell-derived pancreatic progenitor cells from Geusz *et al.* [22]. Of the 1,673 CUT&RUN peaks, 612 overlapped with ATAC-seq peaks in these datasets, indicating that they are likely biologically relevant to beta cell development and function (Supplemental table 3).

**Figure 2:**
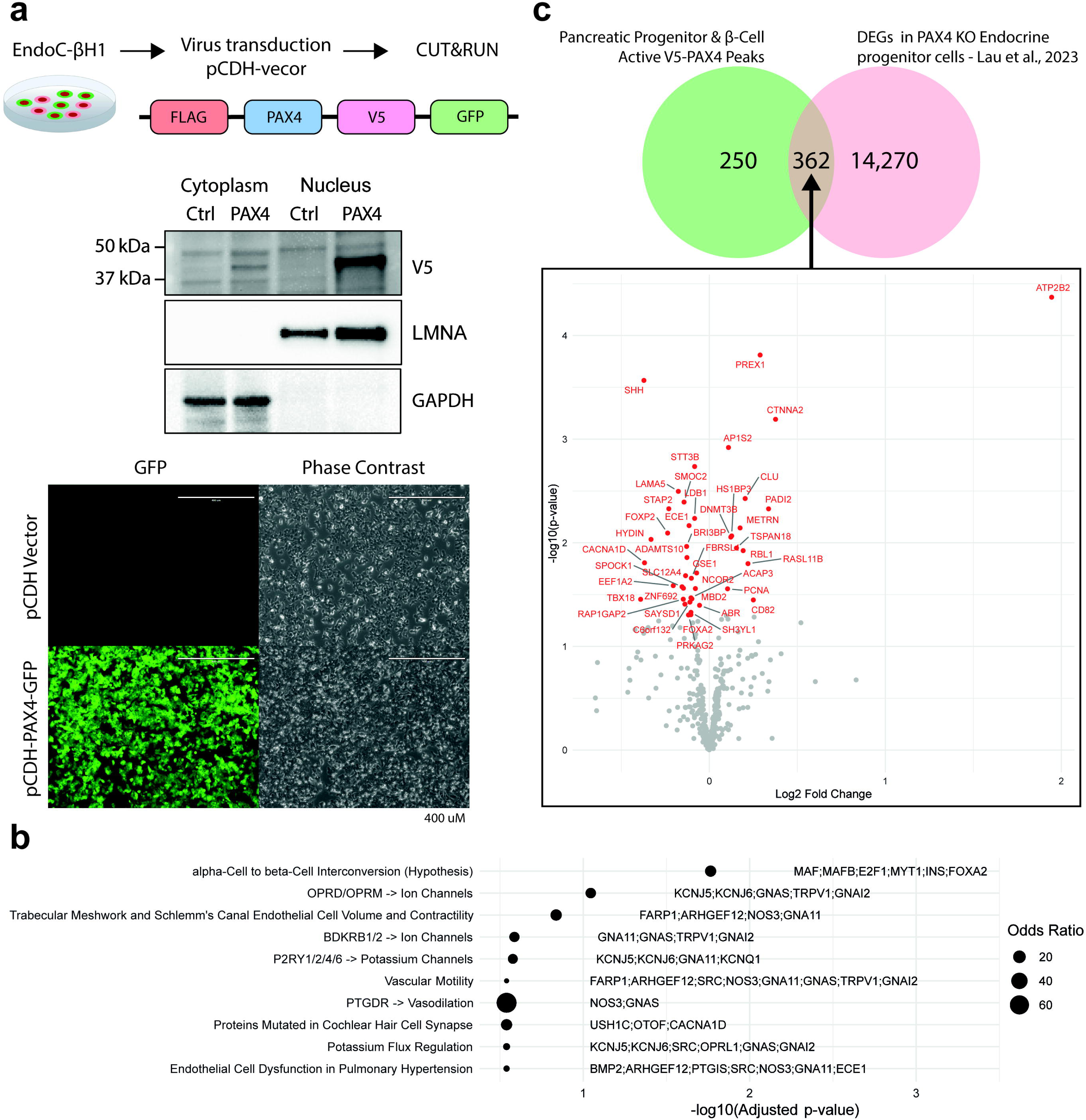
a) Cleavage Under Targets and Release Using Nuclease (CUT&RUN) sequencing of PAX4 binding targets in EndoC-βH1 cells. FLAG-PAX4-V5-GFP was introduced to the cells via lentiviral transduction, with bound regions then sequenced across 5 million reads. Western blotting and fluorescent microscope images confirmed nuclear localization and the overexpression of PAX4 in the EndoC-βH1 cells, respectively. b) 1,673 peaks were identified through analysis with SEACR [20], 612 of which overlapped regions of accessible chromatin in pancreatic progenitors and adult donor beta cells. Gene ontology enrichment analysis revealed a significant enrichment for genes belonging to a hypothesized alpha cell to beta cell interconversion pathway in the Elsevier Pathway Collection among the genes putatively regulated by the 612 accessible PAX4 peaks. c) We compared the genes bound by PAX4 with those that were dysregulated in the Lau *et al.* [12] *PAX4* knockout (KO) in endocrine progenitors, with a total of 362 overlapping genes, 43 of which were significantly differentially expressed in the KO cells compared to the wild-type.

Gene ontology analysis of the genes proximal to the 612 beta cell- and pancreatic progenitor-accessible PAX4 binding peaks in the Elsevier Pathway Collection revealed an enrichment of genes in a hypothesized alpha-cell to beta-cell interconversion pathway (Figure 2b). This was the most significantly enriched pathway in the dataset and the only pathway that was still significant after adjustment for multiple testing (adjusted P = 0.017). The enrichment was driven by the presence of peaks overlapping promoters and enhancers of the genes *MAF*, *MAFB*, *E2F1*, *MYT1*, *INS* and *FOXA2*. The binding of PAX4 to the *INS* gene promoter is direct evidence for the importance of the gene in insulin secretion, while MAFB and FOXA2 are known to have essential roles in islet development and beta cell fate specification from human genetic studies [23,24]. MAF is a known regulator of alpha cell-specific genes and acts cooperatively with PAX6 to transactivate the glucagon promoter [25,26]. E2F1 and MYT1 have documented roles in beta cell proliferation [27,28]. Interestingly, many of the other nominally significantly enriched pathways identified in the analysis were related to potassium ion flux regulation. Potassium ion transportation is vital to insulin secretion[29]; pathogenic activating variants in the *ABCC8* and *KCNJ11* ATP-sensitive potassium channel subunit encoding genes are a common cause of both permanent NDM (PNDM) and TNDM.[15,30]

Comparison of the 612 beta cell- and pancreatic progenitor-accessible peaks with differentially expressed genes (DEGs) in *PAX4* KO stem cell derived endocrine progenitor cells from Lau *et al.* revealed an overlap of 362 genes, 43 of which showed significant differences in expression between the wild-type and KO cells (Figure 2c). The overlap included *FOXA2*, *FOXP2* and *SHH*, which all have well-established essential roles in pancreatic islet development and function [24,31,32]. The overlap also included the *CACNA1D* gene, which encodes a subunit of the Cav1.3 voltage-gated calcium channel that forms part of the glucose-sensitive insulin release pathway [33]. Autosomal dominant activating variants in *CACNA1D* are a known cause of congenital hyperinsulinism, a disease characterized by unregulated insulin secretion from pancreatic beta cells [34]. We also examined whether the PAX4 binding sites identified from the CUT&RUN data are enriched in T2D GWAS loci [35]; however, no significant enrichment was observed. Overall, the analysis of the PAX4 CUT&RUN data shows a clear role of the gene in human islet development and in the regulation of genes important for glucose-sensitive insulin secretion.

### 2.3. Alternative splicing of a 5’ upstream open reading frame is a human-specific regulatory mechanism of PAX4 protein expression

Transcriptomic analysis of *PAX4* highlighted two highly expressed transcripts in developing human pancreatic endocrine cells: the MANE select transcript, and an alternatively spliced version that was not present in either the Ensembl or NCBI RefSeq databases (Figure 3). The alternate transcript was missing the second exon of the 5’ UTR compared to the MANE select transcript and was identified through analysis of RNA-Seq data for differentiating stem cell at the posterior foregut, pancreatic progenitor, endocrine progenitor and islet stages from De Franco *et al.* [5]. This new alternatively spliced transcript was the most highly expressed *PAX4* transcript in all *PAX4*-expressing cell types explored, except endocrine progenitors in which the MANE select showed slightly higher expression. The second non-coding exon missing in the alternate spliced isoform contains a 75bp long 5’ upstream open reading frame (uORF) and thus splicing of this exon may have a role in the regulation of PAX4 protein expression. Ribosomal binding of the uORF was confirmed using Ribo-seq data for pancreatic progenitor cells from Gaertner *et al* [36] (Supplemental figure 1). Previous work in other genes has shown that the presence of such uORFs in coding transcripts can reduce translation of the associated protein by 30-80% [37] and uORFs have been confirmed to play a role in regulating the expression of the closely related PAX6 protein [38]. The alternatively spliced exon is also not present in any of the known mouse *Pax4* transcripts [39] and, as such, could represent a human-specific regulatory mechanism of PAX4 protein expression. As the coding sequence would be unchanged for this novel transcript, we expect that the variants identified in the two individuals with TNDM would have the same effect on this new transcript as had been predicted for the MANE select transcript and result in complete loss-of-function.

**Figure 3:**
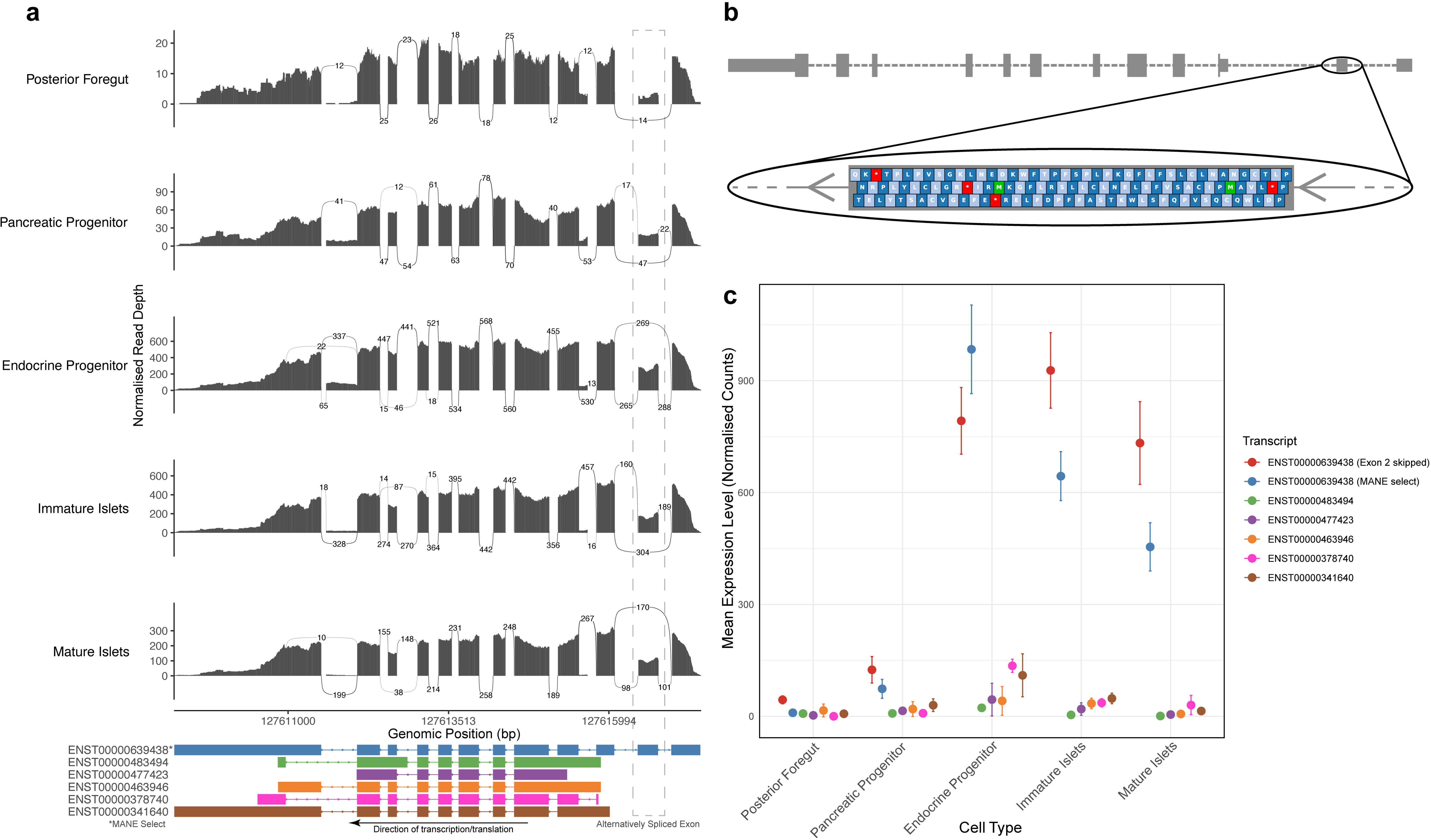
a) Transcript expression of *PAX4* in embryonic stem cell-derived pancreatic islet cells from De Franco *et al.* [5]. Sashimi plots showing exon expression highlight high expression of an alternatively-spliced version of the MANE select transcript where the second, non-coding exon is skipped. b) This exon contains a 75bp long upstream open reading frame. c) Quantification of the known transcripts and this novel transcript show that the novel transcript and the MANE select are the most expressed throughout pancreatic development, with the novel transcript the most expressed in all stages other than endocrine progenitors.

## 3. Conclusion

Our study establishes bi-allelic *PAX4* LoF variants as a novel genetic cause of NDM. The transient diabetes phenotype observed in our patients suggests that beta cells can develop in humans in the absence of PAX4. This differs from observations in rodent *Pax4* KO models that have complete loss of beta cells resulting in permanent neonatal diabetes and postnatal death. Functional profiling of *PAX4* in human pancreatic cell models using CUT&RUN and RNA-Seq highlights this gene’s regulatory role for PAX4 in both islet development and glucose-sensitive insulin secretion. This highlights a specific but non-essential role in human beta-cell development and provides functional evidence for its mechanism not only in monogenic disease but also as a risk gene for type 2 diabetes.

## Acknowledgements

This research was funded in whole, or in part, by the Wellcome Trust [223187/Z/21/Z and 224600/Z/21/Z]. For the purpose of open access, the author has applied a CC BY public copyright license to any Author accepted Manuscript version arising from this submission. This research was supported by the National Institute for Health and Care Research (NIHR) Exeter Biomedical Research Centre (BRC) and National Institute for Health and Care Research Exeter Clinical Research Facility. The views expressed are those of the author(s) and not necessarily those of the NIHR or the Department of Health and Social Care. This research was supported by the Diabetes Research and Wellness Foundation. A.T.H. is employed as a core member of staff within the National Institute for Health Research–funded Exeter Clinical Research Facility and is an NIHR Emeritus Senior Investigator. M.B.J. is funded by a Diabetes UK/Breakthrough T1D RD Lawrence Fellowship. S.E.F. has a Wellcome Trust Senior Research Fellowship [223187/Z/21/Z]. E.D.F. is funded by a Diabetes UK RD Lawrence Fellowship [19/005971]. This work was funded in Stanford by the Wellcome Trust (200837) and the NIDDK (UM1-1DK126185, P30 DK116074; R01 DK140555). The authors acknowledge Dr. Adrian Kee Keong Teo for the gift of the pCDH empty and pCDH-FLAG-PAX4-V5-GFP plasmids.

## CRediT authorship contribution statement

**James Russ-Silsby:** Conceptualization, Data curation, Formal Analysis, Funding acquisition, Investigation, Methodology, Project administration, Software, Validation, Visualization, Writing – original draft, Writing – review & editing; **Yunkyeong Lee:** Conceptualization, Data curation, Formal Analysis, Investigation, Methodology, Project administration, Validation, Visualization, Writing – original draft, Writing – review & editing; **Varsha Rajesh:** Formal Analysis, Investigation, Visualization, Writing – review & editing; **Mahsa Amoli:** Data curation, Resources, Writing – review & editing; **Nasser Ali Mirhosseini:** Data curation, Resources, Writing – review & editing; **Tushar Godbole:** Data curation, Resources, Writing – review & editing; **Matthew B Johnson:** Resources, Writing – review & editing; **Dora E Ibarra:** Methodology, Writing – review & editing; **Han Sun:** Data curation, Formal Analysis, Methodology, Software, Visualization, Writing – original draft, Writing – review & editing; **Nicole A J Krentz:** Conceptualization, Methodology, Writing – review & editing; **Matthew N Wakeling:** Data curation, Methodology, Software, Writing – review & editing; **Sarah E Flanagan:** Resources, Writing – review & editing; **Andrew T Hattersley:** Resources, Writing – review & editing; **Anna L Gloyn:** Conceptualization, Funding acquisition, Methodology, Project administration, Resources, Supervision, Validation, Writing – original draft, Writing – review & editing; **Elisa De Franco:** Conceptualization, Funding acquisition, Methodology, Project administration, Resources, Supervision, Validation, Writing – original draft, Writing – review & editing

## Data Availability

The CUT&RUN Data reported in this study is available through the EGA - https://ega-archive.org[EGAS50000000857]. Applicants should request access to the data using the data access agreement form (available from EGA or through https://med.stanford.edu/genomics-of-diabetes/datasets.html). Upon receipt of completed forms, a decision on access will be made by the data access committee (DAC) within 14 days. The chair of the DAC will inform EGA and EGA will release the data to the account listed in the data access agreement. Forms should be returned to Anna L. Gloyn, Stanford University.

## STAR Methods

### Key Resources Table

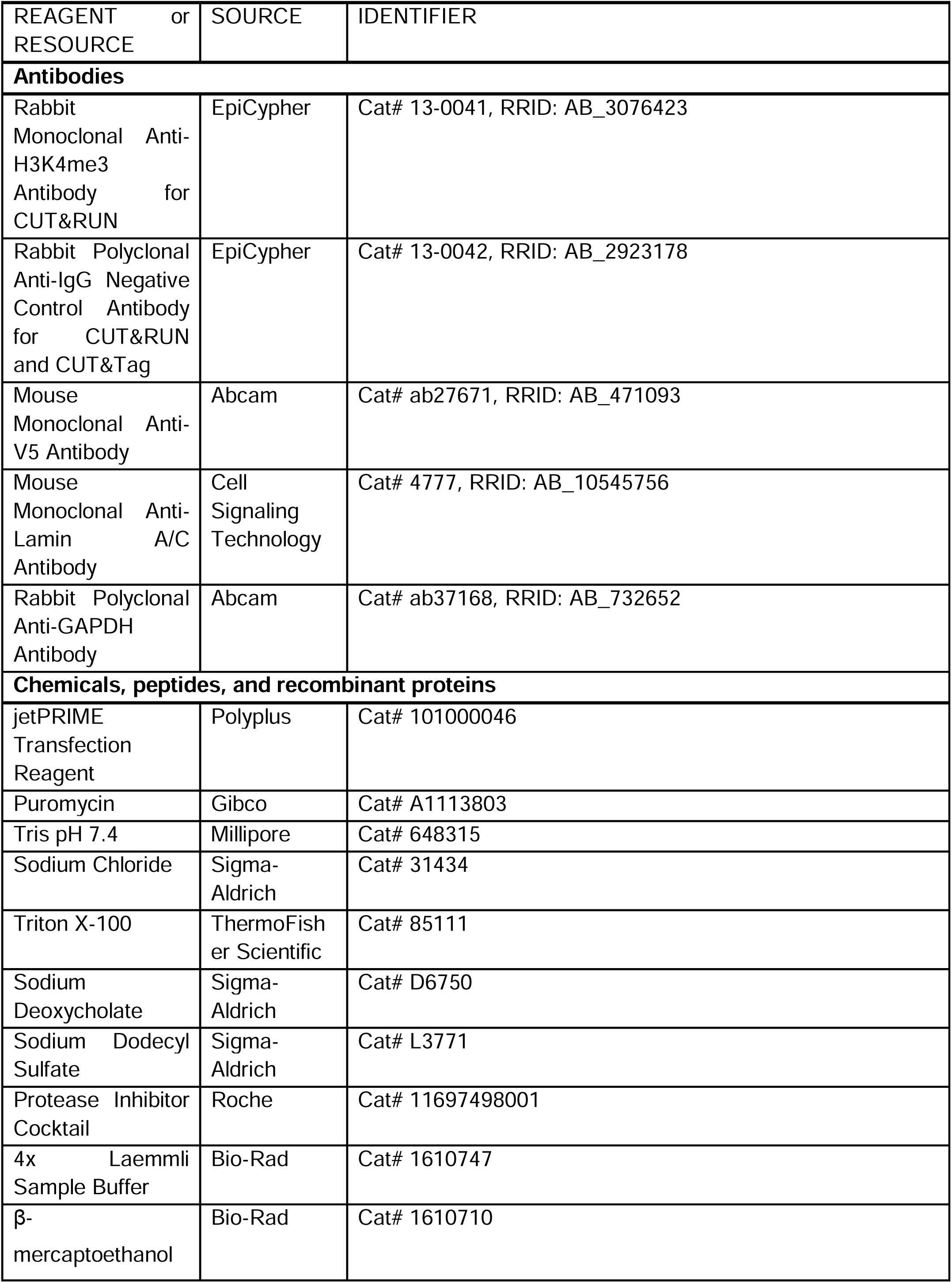
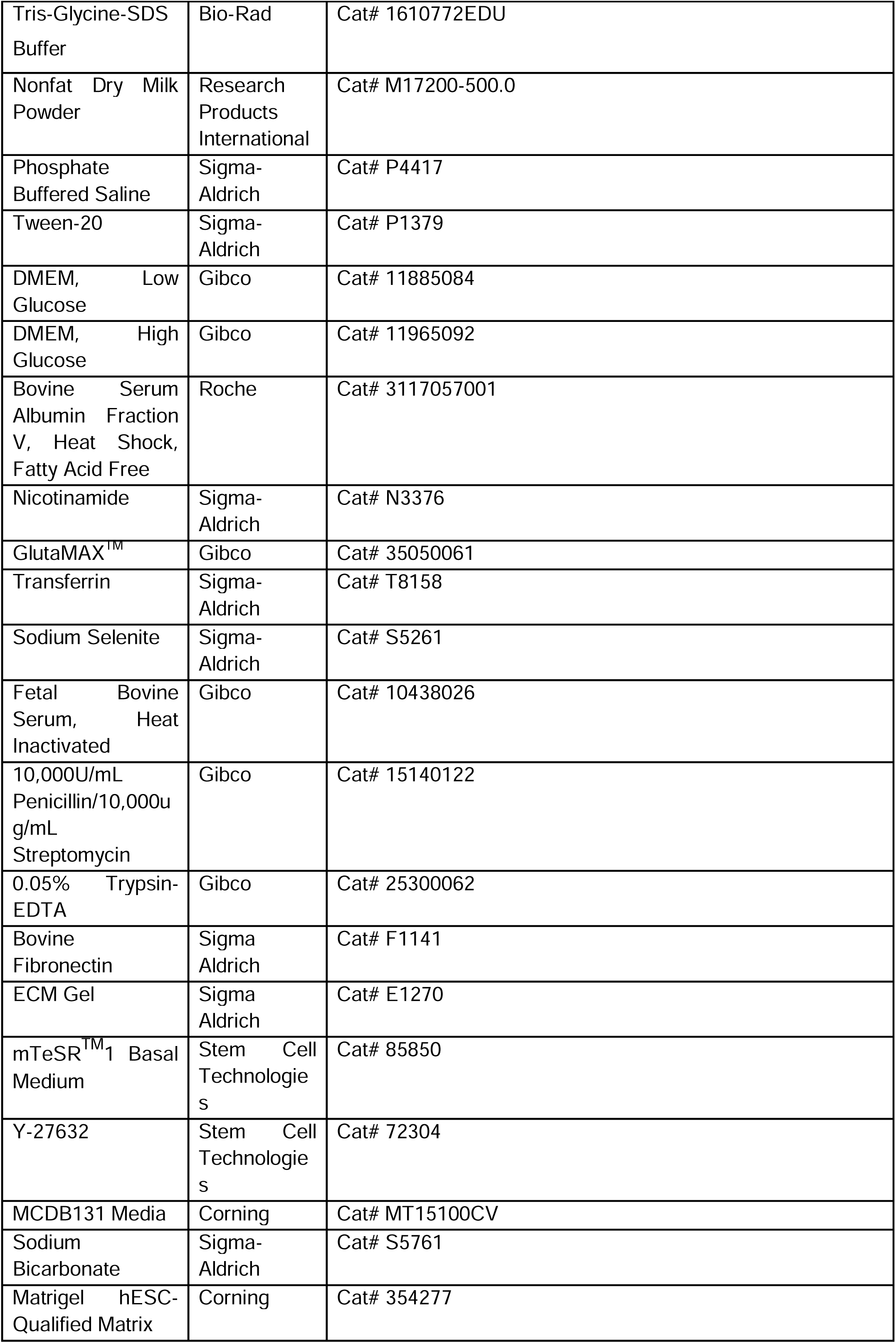
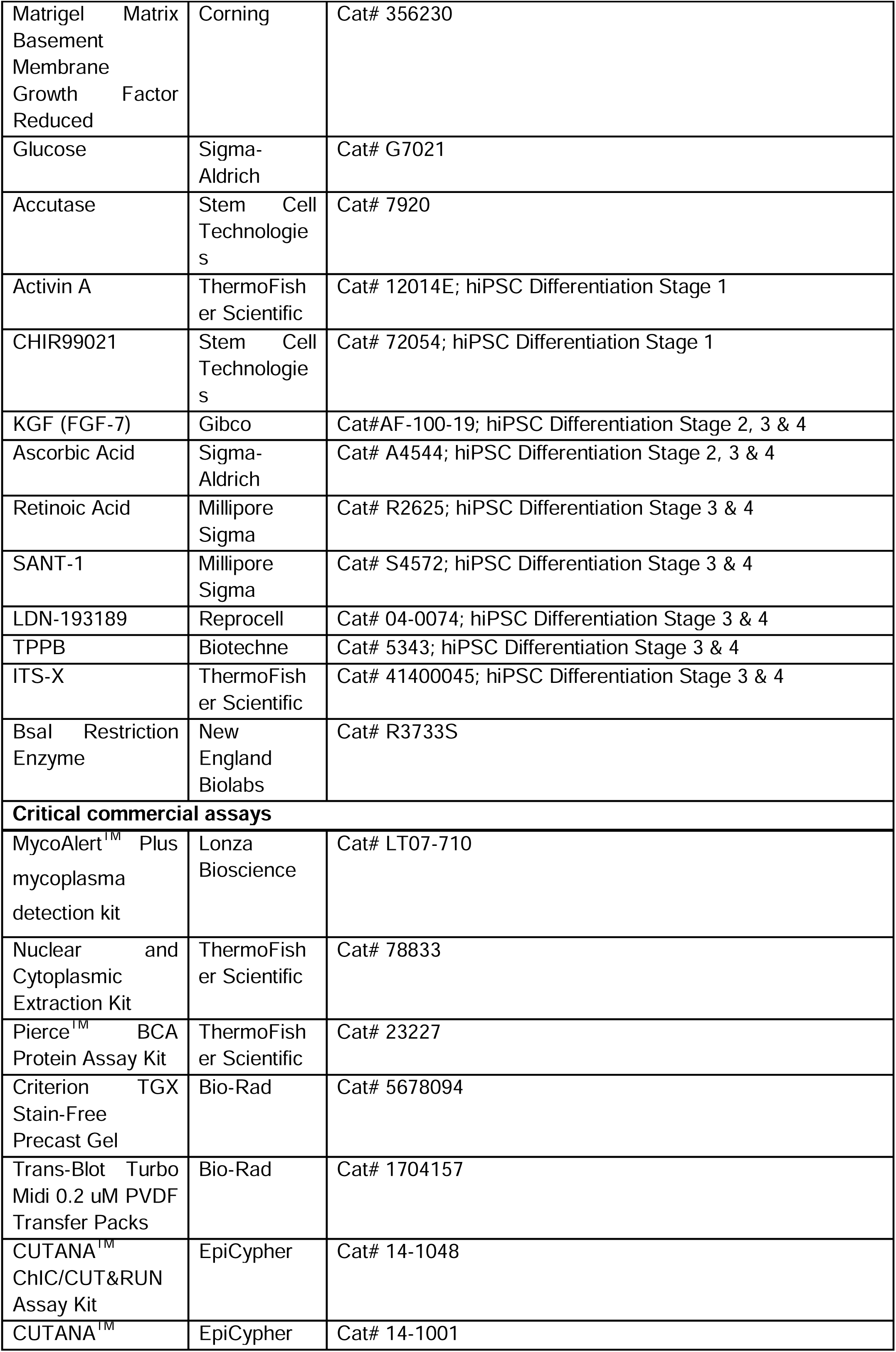
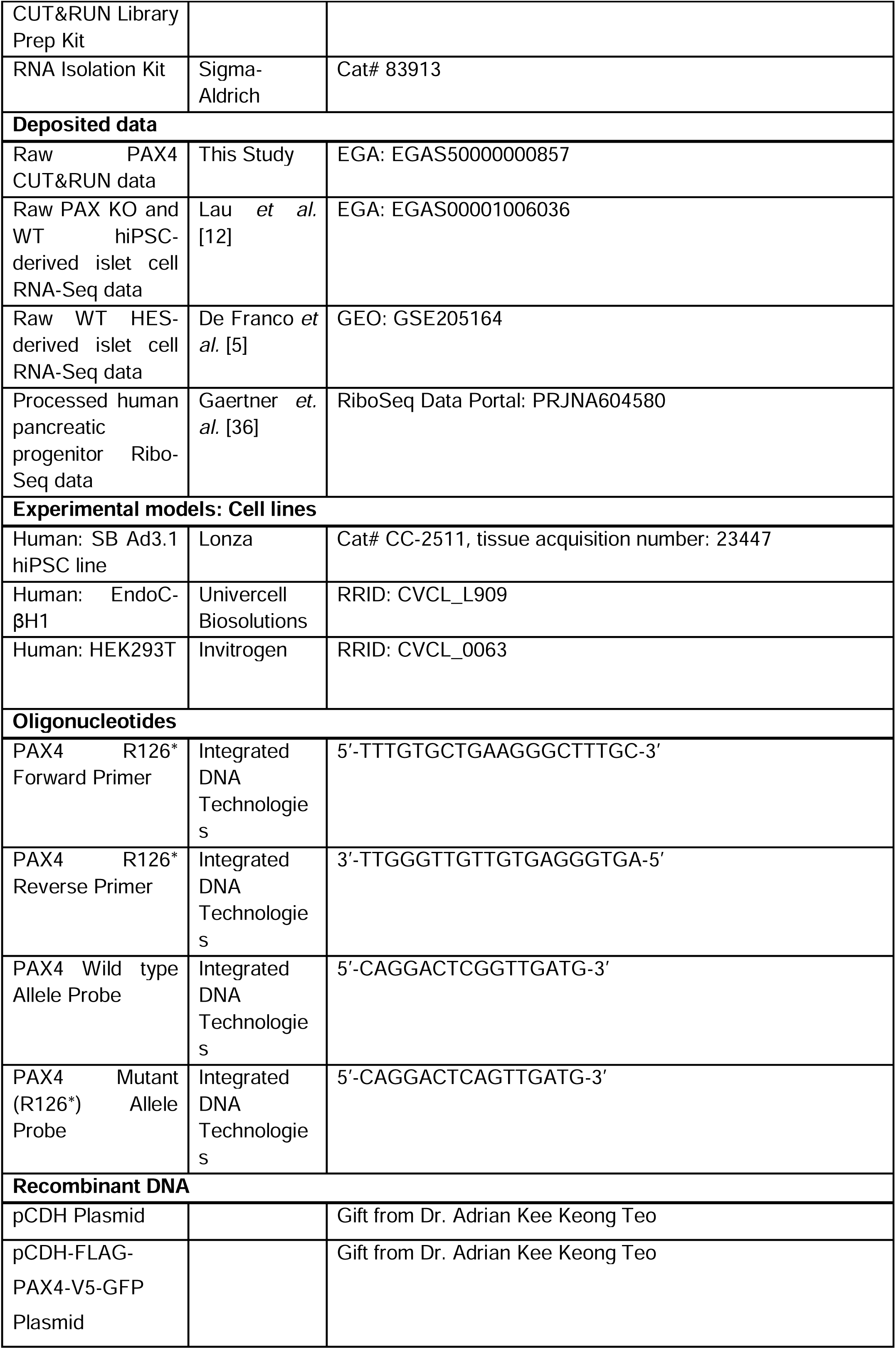
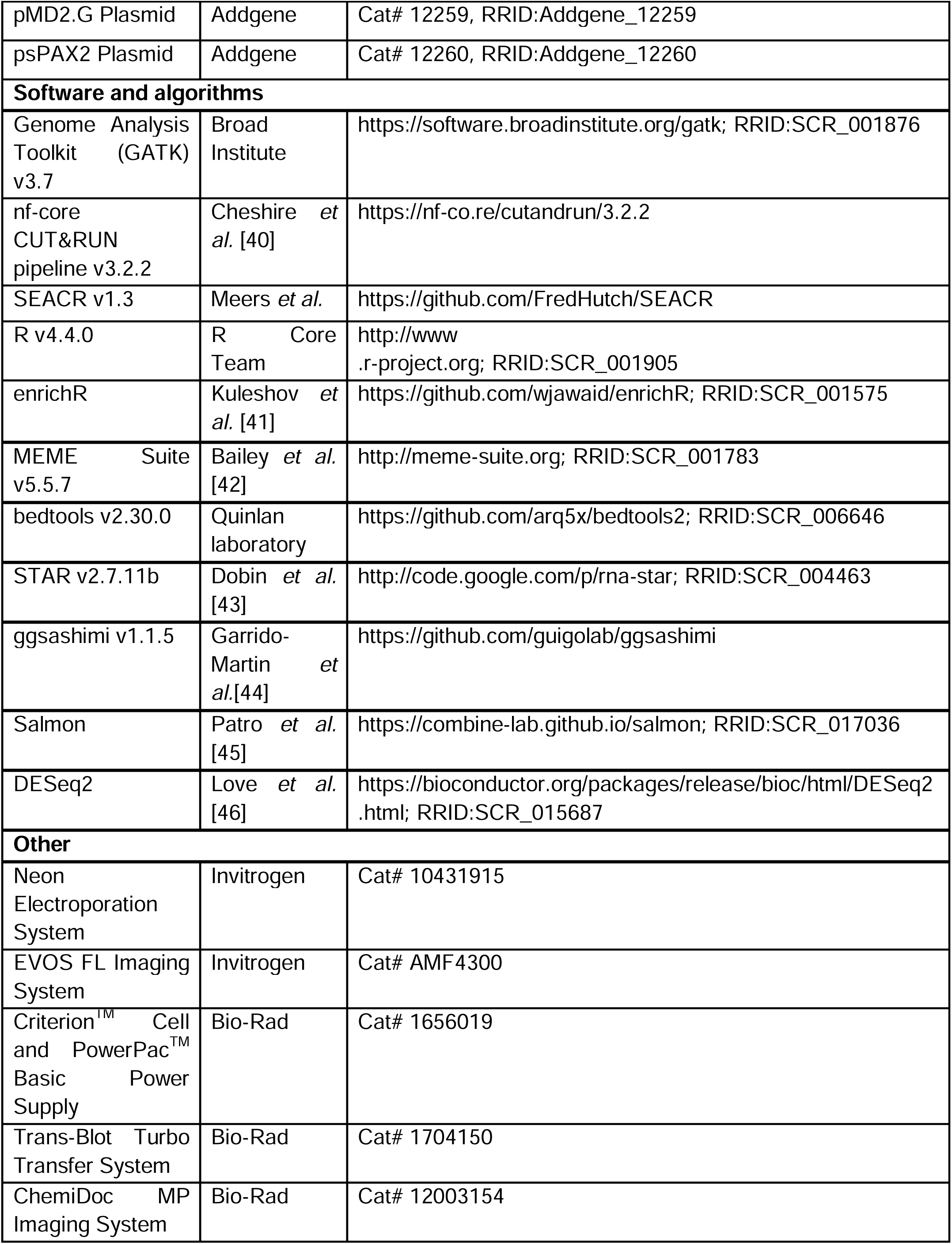

### Experimental model and study participant details

#### Study Participants

Individuals with clinically suspected monogenic diabetes were recruited by their clinicians for molecular genetic analysis in the Exeter Molecular Genetics Laboratory. Consanguineous individuals had either reported parental relatedness at referral or had homozygous segments making up more than 3% of their genome. The study was conducted in accordance with the Declaration of Helsinki, and all subjects or their parents gave informed consent for genetic testing with ethical approval received from the Genetic Βeta-cell Research Bank, Exeter, U.K. (17/WA/0327).

### Human Induced Pluripotent Stem Cells (hiPSCs)-Derived Pancreatic Endoderm (PE) Cells

hiPSCs were cultured on hESC-qualified matrigel (Corning, 354277)-coated plates in mTeSR^TM^1 basal medium (STEMCELL^TM^ Technologies, 85850) supplemented with 1% Penicillin/Streptomycin (Gibco, 15140122) and 10 uM ROCK inhibitor (Y-27632, STEMCELL^TM^ Technologies, 72304) with daily media changes and passaged using Accutase (STEMCELL^TM^ Technologies, 7920). A 200 nucleotide single-stranded oligodeoxynucleotide repair template (purchased from Integrated DNA Technologies) was designed containing the p.(Arg126*) (Chr7(GRCh37):g.127254596G>A) variant. A silent mutation was also included at codon 126 to remove BsaI/BseRI restriction enzyme sites for genotyping. 1 x 10^6^ NKX6.1-GFP hiPSCs[47] were electroporated with an RNP complex (gRNA, Alt-R Cas9 enzyme) and Alt-R HDR donor oligo using the Neon Electroporation System (Invitrogen, 10431915). Electroporate using following parameters: 1,200 volts, 30 ms, 1 pulse. Single-cell colonies were picked, expanded and screened using the BsaI restriction enzyme (New England Biolabs, R3733S). All clones (p.(Arg126*) WT sham, Het and Hom) were further validated by Sanger sequencing using the 5’-TTTGTGCTGAAGGGCTTTGC-3’ (forward) and 3’-TTGGGTTGTTGTGAGGGTGA-5’ (reverse) primer sequences. CRISPR genome edited hiPSCs were seeded on Growth Factor Reduced Basement Membrane Matrigel (Corning, 356230)-coated plates in mTeSR^TM^1 basal medium supplemented with 10 uM ROCK inhibitor. The differentiation was started following day of seeding in MCBD131 media (Corning, MT15100CV) supplemented 1% Penicillin/Streptomycin, sodium bicarbonate (Sigma-aldrich, S5761), GlutaMAX^TM^ (Gibco, 35050061) and glucose (Sigma-aldrich, G7021) and proceeded through a seven-stage differentiation protocol (stages 1-4; stage 1: definitive endoderm (DE), stage 2: primitive gut tube (PGT), stage 3: posterior foregut (PFG) and stage 4: pancreatic endoderm (PE)) [14].

### EndoC-**β**H1 Cells

EndoC-βH1 cells were cultured in low glucose DMEM (Gibco, 11885084) supplemented with 2% BSA (Roche, 3117057001), 10 mM nicotinamide (Sigma-Aldrich, N3376), 2 mM GlutaMAX^TM^ (Gibco, 35050061), 50 μM β-mercaptoethanol (Bio-Rad, 1610710), 5.5 μg/mL transferrin (Sigma-Aldrich, T8158), 6.6 ng/mL sodium selenite (Sigma-Aldrich, S5261) and 1% Penicillin/Streptomycin (Gibco, 15140122) in tissue culture plates precoated with high glucose DMEM (Gibco, 11965092) supplemented with 2 μg/mL bovine fibronectin (Sigma-Aldrich, F1141) and 1% ECM (Sigma-Aldrich, E1270) in a 5% CO_2_ humidified incubator at 37°C. Cells were passaged weekly with 0.05% Trypsin/EDTA (Gibco, 25300062).

### HEK293T Cells

HEK293T cells were cultured in high glucose DMEM (Gibco, 11965092) supplemented with 10% Fetal Bovine Serum (Gibco, 10438026) and 1% Penicillin/Streptomycin (Gibco, 15140122). Cells were passaged twice a week with 0.05% Trypsin/EDTA. All mammalian cells were regularly tested to be mycoplasma free using a MycoAlert^TM^ Plus mycoplasma detection kit (Lonza Bioscience, LT07-710).

## Method Details

### Genome Sequencing

DNA was extracted from peripheral blood mononuclear cells and saliva samples and sequenced using an Illumina HiSeq 2500, an Illumina HiSeq X10 and a BGI-Seq 500, producing 100bp and 150bp paired-end reads. Genome sequencing data was then processed according to the GATK best-practice workflow for genome sequencing data (https://gatk.broadinstitute.org).

### Targeted Next Generation Sequencing

Target regions were captured and enriched using Agilent SureSelect custom capture and Twist Bioscience Custom panel kits. These regions were then sequenced using an Illumina NextSeq or an Illumina NovaSeq X, producing 150bp paired-end reads. The sequencing data was processed using a GATK best-practice workflow optimized for targeted and exome sequencing (https://gatk.broadinstitute.org).

### Allele-Specific qPCR of CRISPR Edited Human Induced Pluripotent Stem Cell (iPSC)-Derived Pancreatic Endoderm Cells

Pancreatic endoderm samples were lysed and RNA extracted using RNA isolation kit (Sigma-Aldrich, 83913). RNA was treated with DNase to remove any genomic DNA and quality and concentration determined on Nanodrop. mRNA was reverse transcribed to cDNA. cDNA was then incubated with a SNP genotyping assay containing two probes that separately bound the WT and stop-gain p.(Arg126*) *PAX4* alleles. Both probes were coupled with unique fluorophores to separately track amplification of the 2 alleles with each round of PCR (HEX and FAM).

### CUT&RUN Assay, Library Preparation and Sequencing in EndoC-βH1 cells

CUT&RUN was performed on nuclei extracts from FLAG-PAX4-V5-GFP overexpressing EndoC-βH1 cells using CUTANA^TM^ ChIC/CUT&RUN Kit version 2.1 (EpiCypher, 14-1048) according to the manufacturer’s instructions. Briefly, samples were incubated with 0.5 ug of mouse anti-V5 tag (Abcam, ab27671), H3K4me3 positive control (EpiCypher, 13-0041) and rabbit anti-IgG negative control (EpiCypher, 13-0042) antibodies. Library preparation was performed using the CUTANA^TM^ CUT&RUN Library Prep Kit (version 1.2, EpiCypher, 14-1001). Libraries were sequenced using Novaseq 5M paired end reads per sample.

### Lentiviral Transduction and Overexpression in EndoC-**β**H1

pCDH empty and pCDH-FLAG-PAX4-V5-GFP plasmids [12] were co-transfected in HEK293T with packaging vectors pMD2.G (Addgene, 12259) and psPAX2 (Addgene, 12260) using jetPRIME transfection reagent (Polyplus, 101000046). Viral supernatant was collected at 72h post-transfection and EndoC-βH1 cells were transduced overnight and selected for 7d in 4 μg ml^-1^ puromycin (Gibco, A1113803) with media changes as required. Overexpression of the GFP including construct was confirmed using the EVOS FL Imaging System (Invitrogen, AMF4300).

### Nuclear and Cytoplasmic Extraction and Western Blot Analysis

Nuclear and Cytoplasmic Extraction Kit (ThermoFisher, 78833) was used for cell lysis and extraction of separate nuclear and cytoplasmic protein fractions according to the manufacturer’s specifications. For SDS-PAGE, whole cell protein lysates were obtained from cell pellets through lysis in RIPA buffer (50 mM Tris pH 7.4 (Millipore, 648315), 150 mM sodium chloride (Sigma-Aldrich, 31434), 1% Triton X-100 (ThermoFisher Scientific, 85111), 0.5% sodium deoxycholate (Sigma-Aldrich, D6750), 0.1% sodium dodecyl sulfate (Sigma-Aldrich, L3771)) containing 1x protease inhibitor cocktail (Roche, 11697498001). Protein was quantified using Pierce^TM^ BCA Protein Assay Kit (ThermoFisher Scientific, 23227). 15 ug of total protein in 4x Laemmli sample buffer (Bio-Rad, 1610747) containing 10% β-mercaptoethanol (Bio-Rad, 1610710), and boiled for 5 min at 100°C. Denatured samples were separated on 4-20% Criterion^TM^ TGX Stain-Free Precast Gel (Bio-Rad, 5678094) in Tris-glycine-SDS buffer (Bio-Rad, 1610772EDU) using Criterion^TM^ Cell and PowerPac^TM^ Basic Power Supply (Bio-Rad, 1656019) and the gel was transferred onto a 0.2 μM polyvinylidene difluoride (PVDF, Bio-Rad, 1704157) membrane using the Trans-Blot Turbo Transfer System (Bio-Rad, 1704150). Nonspecific antibody binding was blocked in 5% dry milk powder (Research Products International, M17200-500.0) in 1x phosphate buffered saline (Sigma-Aldrich, P4417) containing 0.1% Tween-20 (Sigma-Aldrich, P1379) (PBST) and the membranes were incubated with primary antibodies (V5 (Abcam, ab27671), LMNA (Cell Signaling Technology, 4777) and GAPDH (Abcam, ab37168)) at 4°C overnight and secondary antibodies at room temperature for 1h. Blots were acquired and quantified using the ChemiDoc MP Imaging System (Bio-Rad, 12003154).

### Quantification and statistical analysis

#### CUT&RUN Analysis

CUT&RUN Fastq files from the CUT&RUN experiments were processed following the nf-core CUT&RUN pipeline (https://nf-co.re/cutandrun/3.2.2/) [40]. Peaks were called using the SEACR peak caller under the stringent parameter [20]. ATAC-Seq peaks in adult donor beta cells from Chiou *et al.*[21] and pancreatic progenitor cells from Geusz *et al.* [22] were used to prioritize binding sites that were likely to be functionally relevant. Gene ontology analysis was performed using EnrichR (https://github.com/wjawaid/enrichR) [41] in R based on the Elsevier Pathway Collection. To assess whether the PAX4 binding sites identified from the CUT&RUN data are enriched in T2D GWAS loci, we downloaded the credible sets from the Mahajan et al. [35] multi-ancestry genetic study, converted the coordinates from hg19 to hg38, and performed a Fisher’s exact test using bedtools (v2.30.0). Find Individual Motif Occurrences (FIMO) and Multiple Em for Motif Elicitation (MEME) from the MEME Suite v5.5.7 [42] were also used to examine motif enrichment within the CUT&RUN peaks (supplemental table 4, supplemental figure 2).

### Transcriptomic Analysis in Stem Cell-Derived Islets

RNA-Seq data for PAX4 knockout (KO) and corresponding wild type (WT) induced pluripotent stem cell (iPSC) derived endocrine progenitor cells from Lau *et al.* [12] (protocol B) was analysed as previously described to identify differently expressed genes in the KO cells.

RNA-Seq data for embryonic stem cell-derived posterior foregut, pancreatic progenitor, endocrine progenitor, immature islet and mature islet cells that was generated as part of De Franco *et al.* [5] was aligned to the GRCh38 reference using STAR [43]. Sashimi plots were generated using ggsashimi [44]. Transcripts were then quantified at each stage using Salmon[45] and DESeq2 [46] using a custom .gtf reference that included the newly identified exon skipped transcript.

### Ribo-Seq Data Analysis

Pre-processed pancreatic progenitor Ribo-seq data from Gaertner *et. al.* [36] was downloaded from RiboSeq.org [48] in bigwig format. The data was then re-aligned to the PAX4 MANE-select transcript using R and visually inspected for ribosomal binding peaks overlapping the uORF identified in the 2nd exon.

