## Supplemental Figure 1,2 and Supplemental Table 1 for "Complete Loss of PAX4 causes Transient Neonatal Diabetes in Humans"

### Supplemental Figures

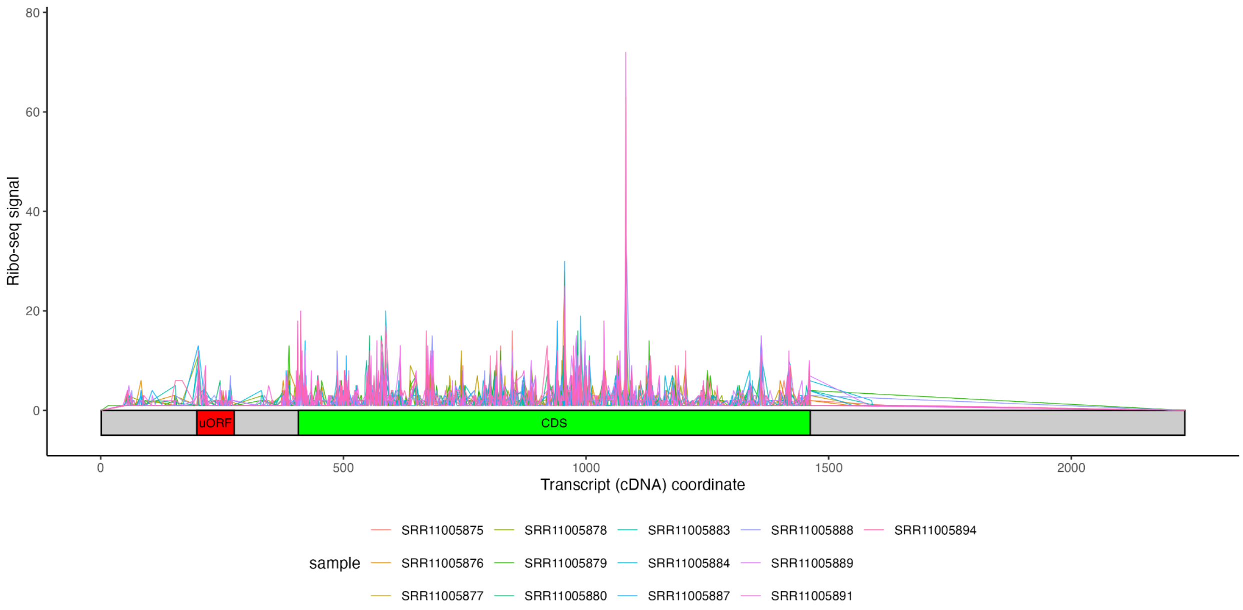


Supplemental figure 1: Pancreatic progenitor Ribo-seq data from Gaertner *et al.* aligned to the PAX4 MANE Select cDNA sequence. The presence of a peak corresponding to the uORF confirms it is being bound by the ribosome during PAX4 protein translation.

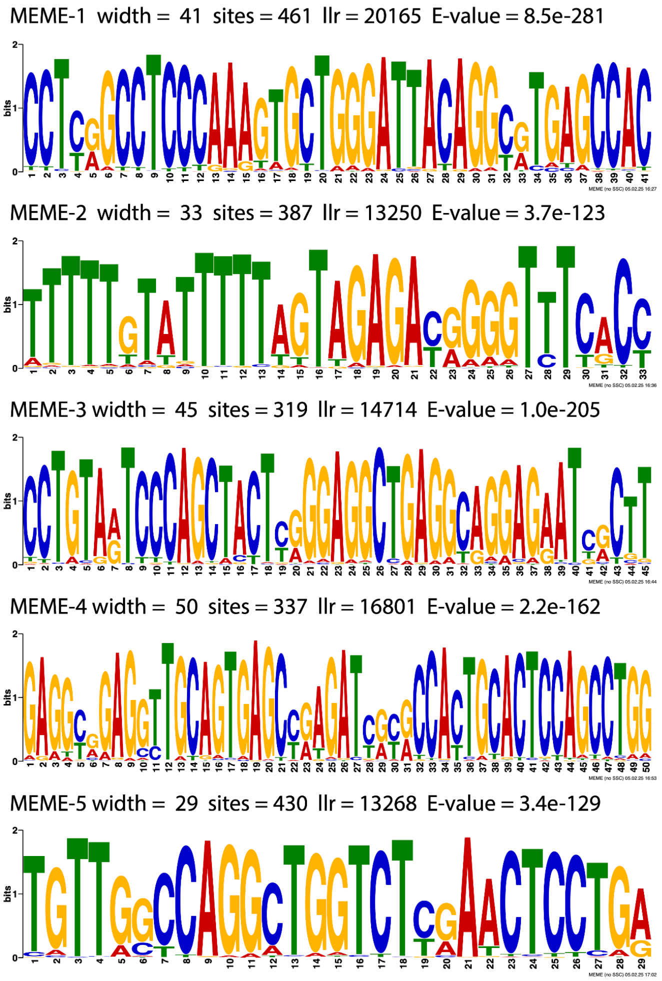


Supplemental figure 2: Motifs discovered using MEME on the PAX4 CUT&RUN peaks. Multiple Em for Motif Elicitation (MEME) from the MEME Suite (v5.5.7) was used to discover novel motifs from the genomic sequences of the peaks identified in the CUT&RUN data. The top 5 motifs discovered are reported here.

**Supplemental Tables**

Supplemental table 1: Clinical features of patients with biallelic *PAX4* variants

| | **Individual 1 (I-1)** | **Individual 2 (I-2)** |
|---|---|---|
| **Homozygous *PAX4* Variant** | p.(Arg126*) | c.-352_104del |
| **Birth weight** | -1.16SD | -2.98SD |
| **Diabetes Diagnosis Age** | 1-5 months (diabetic ketoacidosis) | 1-5 months (diabetic ketoacidosis) |
| **Diabetes Remission Age** | 6-12 months | 6-12 months |
| **Diabetes Relapse Age** | 2-7 years | 2-7 years |
| **Current treatment** | Basal Bolus Insulin (1U/Kg/Day) + Glibenclamide | Basal Bolus Insulin (Dose Unknown) |
| **Additional features** | Mild Learning Disorder | |
| **Random non-fasting C-peptide (Age)** | Unknown | 1ng/ml (2-7 Years) |
| **Most Recent HbA1c (Age)** | 53 mmol/mol (8-13 Years) | 79 mmol/mol (2-7 Years) |
